# Attitudes and Perceptions Toward the Use of Artificial Intelligence Chatbots for Peer Review in Medical Journals: A Large-Scale, International Cross-Sectional Survey

**DOI:** 10.64898/2026.04.07.26350263

**Authors:** Jeremy Y. Ng, Daivat Bhavsar, Neha Dhanvanthry, Lex Bouter, Teresa Chan, Holger Cramer, Annette Flanagin, Alfonso Iorio, Cynthia Lokker, Hervé Maisonneuve, Ana Marušić, David Moher

## Abstract

**Background:** Artificial intelligence chatbots (AICs), as a form of generative artificial intelligence (AI), are increasingly being considered for use in scholarly peer review to assist with tasks such as identifying methodological issues, verifying references, and improving language clarity. Despite these potential benefits, concerns remain regarding their reliability, ethical implications, and transparency. Evidence on how medical journal peer reviewers perceive the role and impact of AICs is limited. This study explored reviewers’ familiarity with AICs, perceived benefits and challenges, ethical concerns, and anticipated future roles in peer review.

**Methods:** We conducted a cross-sectional online survey of medical journal peer reviewers. Corresponding author information was extracted from MEDLINE-indexed articles added to PubMed within a two-month period using an R-based approach. A total of 72,851 authors were invited via email to participate; those who self-identified as peer reviewers were eligible. The 29-item survey assessed familiarity with AICs and perceptions of their benefits and limitations in peer review. The survey was administered via SurveyMonkey from April 28 to June 16, 2025, with two reminder emails sent during the data collection period.

**Results:** A total of 1,260 respondents completed the survey. Most participants were familiar with AICs (86.2%) and had used tools such as ChatGPT for general purposes (87.7%), but the majority had not used AICs for peer review (70.3%). Most respondents reported that their institutions do not provide training on AIC use in peer review (69.5%), although many expressed interest in such training (60.7%). Perceptions of AIC benefits were mixed, while concerns were widely shared, particularly regarding potential algorithmic bias (80.3%) and issues related to trust and user acceptance (73.3%).

**Conclusions:** While familiarity with AICs is high among medical journal peer reviewers, their use in peer review remains limited. There is clear interest in training and guidance, however, concerns related to ethics, data privacy, and research integrity persist and should be addressed before broader implementation.

## Background

Artificial intelligence (AI) broadly refers to the capability of computer systems or computer-controlled robots to perform tasks typically associated with human intelligence, such as reasoning, problem-solving, generalizing, and learning from past experiences [1, 2]. While current AI programs lack the versatility of human intelligence, specialized applications have permeated numerous fields, including self-driving cars, speech transcription, healthcare, and education [2]. In many domains, AI has demonstrated benefits such as increased productivity, reduced errors, and cost savings. For example, in the field of medicine, AI systems can improve diagnostic accuracy, optimize treatment plans, and reduce healthcare costs for many healthcare systems [3, 4]. These realized and hypothetical benefits have sparked interest in applying AI to scholarly publishing, including the peer review process. Peer review in the context of medical scholarly publishing refers to a quality control protocol whereby methodology and content experts are entrusted with reviewing and appraising submitted manuscripts [5]. The “peers” provide arms-length assessments and constructive feedback to uphold the validity and integrity of manuscript content prior to potential dissemination, in accordance with the varying responsibilities and requirements set by the academic publisher or individual journal [5]. Although this remains a promising measure, improvements in both the efficiency and the quality of peer review are very much needed [5].

Artificial intelligence chatbots (AICs), a subset of AI programs, are generative AI tools designed to simulate human conversation via text or speech [6]. AICs have demonstrated versatility in applications ranging from customer service to education [6, 7]. Within scholarly publishing, AICs hold promise for tasks such as improving language clarity, identifying methodological flaws, verifying references, and potentially standardizing review quality [8]. Their integration into peer review workflows could potentially alleviate reviewer workloads, streamline processes, and address longstanding issues such as bias and inconsistency between review reports [8]. Moreover, AICs may address reviewer fatigue and help peer reviewers navigate the increasing volume of requests (e.g., manuscript submissions), ensuring more timely and consistent evaluations [9, 10].

Despite these potential advantages, major concerns include the reliability of AICs in evaluating complex scientific content, the risk of overlooking critical nuances, the potential for manipulation within the peer review process, and ethical considerations regarding their use [11]. For example, well-documented limitations of ChatGPT include generating plausible though incorrect or misleading content (i.e., ‘hallucinating’), citation inaccuracies, and referencing non-existent sources [11, 12, 13]. Additionally, as AICs rely on training datasets that may lack novelty or inclusivity, there is a risk of perpetuating outdated information and biases [14].

Ethical concerns also extend to issues of plagiarism, confidentiality, intellectual property rights, transparency, research integrity, and accountability [8]. While the use of AICs in peer review may be acceptable under open peer review models that require authors to consent to the broader sharing of manuscript content, this practice raises ethical and procedural concerns for the peer review of manuscripts received by journals that operate closed (i.e., confidential) peer review models [15]. In such cases, sharing unpublished manuscripts with AICs, particularly those hosted on proprietary platforms that store user input, could compromise reviewer confidentiality and violate copyright and other journal policies [15]. Furthermore, for certain medical articles, reviewers may have access to sensitive patient information that is not intended for publication. Sharing this information with AI systems may constitute a breach of patient privacy and, in some jurisdictions, such as the United States, could potentially violate the Health Insurance Portability and Accountability Act (HIPAA) [16, 17].

The scholarly publishing community has started to address these issues through policies for editors and peer reviewers [18–22]. Academic publishers, such as *Nature, JAMA Network,* and *PLOS*, have incorporated guidelines on the use of AICs in their editorial processes, requesting that peer reviewers refrain from uploading any manuscript information to AICs, and that any use of AI for peer review (i.e., evaluation of manuscript claims) must be disclosed in review forms [18–20]. Other organizations, including the World Association of Medical Editors (WAME) and the International Committee of Medical Journal Editors (ICMJE) have recommended that the use of AICs should be prohibited in cases where confidentiality cannot be guaranteed, and if reviewers do use AICs, they must have permission from the journal with clear disclosure of how they were used [23, 24]. However, codifying and regulating the responsible use, or the lack thereof, by peer reviewers may become increasingly difficult with the rapidly evolving possibilities and popularity of AICs.

Despite growing interest in the role of AICs in peer review, there remains a scarcity of available research on the attitudes and perceptions of peer reviewers toward their use [25]. As interest in using AI within the domain of scholarly publishing continues to rise, it is crucial to understand how peer reviewers perceive and engage with AICs, along with the impacts they foresee on the peer review process and publishing overall. This understanding will help leverage the potential of AICs while mitigating its limitations, ultimately helping to enhance the quality, transparency, and fairness of the peer review process. Our study addressed this gap by conducting an international cross-sectional survey to assess medical journal peer reviewers’ familiarity or experience with using AICs, and their perceived benefits and challenges, ethical concerns, and anticipated roles for AICs in the peer review process. By providing insights into reviewer perspectives, this study aims to inform the development of ethical guidelines, practical recommendations, and evaluations needed for the possible effective integration of AICs in peer review.

## Methods

### Open Science Statement

A complete study protocol, with the data analysis plan has been registered on the Open Science Framework (OSF) (https://doi.org/10.17605/OSF.IO/FHC2M) prior to participant recruitment. All relevant study materials and deidentified data can also be found on OSF (https://doi.org/10.17605/OSF.IO/VARJE).

### Ethical Considerations

Prior to conducting this study, ethics approval was obtained from the University Hospital Tübingen Research Ethics Board (REB) (REB Number: 080/2025BO2). Informed consent was obtained from all respondents prior to their participation in the survey, as detailed in the email invitation. Participation in the survey was voluntary, and participants had the right to withdraw from the study at any point prior to submitting their completed survey. All data were confidential and anonymous, and no identifying information of respondents was collected.

### Study Design

We conducted a large-scale, online, closed cross-sectional survey of medical journal peer reviewers, specifically, corresponding authors of articles published in MEDLINE-indexed journals who self-identified as peer reviewers, to investigate their attitudes and perceptions toward the use of AICs in the peer review process.

### Sampling Framework

A comprehensive list of all journals indexed in MEDLINE (approximately 5300 as of November 2024) was gathered along with their NLM IDs [26]. A search strategy, developed by JYN and reviewed by the rest of the team, of the NLM IDs was executed in OVID MEDLINE (**Appendix 1**, https://osf.io/varje/files/ncajv). The search was limited to records indexed over the two months prior to searching (2024/09/01-2024/10/15).

This time frame was selected because researchers who have published within this period were likely to still be actively engaged in research and providing peer review within their fields and available to respond to the survey invitation. Corresponding authors of all types of research articles were considered for inclusion. For articles with two or more corresponding authors, each author was sent an invitation.

Duplicate records were removed prior to the recruitment process. PubMed Identifier (PMID) numbers associated with all yielded articles were exported from OVID as a .csv file and inputted into an R script (created based on the easyPubMed package (easyPubMed, 2023) to capture author names, affiliated institutions, and email addresses from PubMed [27]. A total of 121,995 PMIDs were retrieved and exported into R [27]. Corresponding author name and email address extraction was completed on December 27, 2024. All generated results were combined into a master list and cleaned for potential errors or duplicates before survey administration.

### Inclusion Criteria

To be eligible for participation, participants must have self-identified as medical researchers and served as a peer reviewer of research articles submitted to medical journals. Eligible participants must have completed and submitted at least one peer review report to at least one professional-level medical journal within the past 24 months. Those who had peer reviewed exclusively for student journals (e.g., high school, undergraduate or graduate journals) were not eligible to participate.

### Participant Recruitment

As PMIDs were collected from all MEDLINE-indexed journals, a range of academic disciplines within the field of medicine were included (e.g., public health, bioengineering, medical education). We used convenience sampling to recruit respondents, targeting corresponding authors identified by our sampling strategy. An email was sent to potential participants containing a recruitment message that outlined the purpose of the study and included a link to the survey, using SurveyMonkey, a secure online survey platform (https://www.surveymonkey.com/) [28]. Upon clicking the survey link, participants were directed to a webpage with an informed consent form. Participants had to indicate consent on the form to proceed to the survey. This was a closed survey; only invited participants were able to participate.

After deduplicating the list, a total of 72,851 corresponding authors were sent invitations to participate, in batches of 10,000. If participants did not respond to the initial invitation email sent on April 28, 2025, reminder emails were sent twice during the weeks of May 5, 2025, and May 12, 2025. A 5-week waiting period followed the final email reminder to accommodate remaining interested respondents before the survey was closed permanently. Overall, participants had a total of seven weeks to complete the survey which was closed on June 16, 2025.

No monetary compensation was offered to participants, and participation was completely voluntary and anonymous. Participants could skip the questions that they did not wish to answer and could withdraw from the survey at any time by simply closing the browser window. Participation was anonymous and confidential throughout the study; however, settings allowed for tracking of responded participants (i.e., submission status based on their individual email invitation) for purposes of follow-up reminders and limiting one response per participant.

Withdrawal was not possible after the survey was submitted given that the responses were collected anonymously. Email addresses of all invitees were removed from the mailing system following the 7-week window.

### Survey

The complete survey can be found in **Appendix 2** (https://osf.io/varje/files/c748y). The survey was created, distributed, and collected using SurveyMonkey (Symphony Technology Group, San Mateo, California, United States of America, https://www.surveymonkey.com/). The first draft of the survey was created by JYN following a review of the literature and input from experts in AI and peer review. The survey was then reviewed by all authors. All authors of the study protocol reviewed the survey draft, and the survey was piloted with 5 peer reviewers, exclusive of the author list, to ensure clarity, relevance, and comprehensiveness of the questions prior to circulation. The invited pilot testers were excluded from the final survey.

The survey consisted of 29 questions which included both closed-ended questions (e.g., multiple choice, yes/no) and open-ended questions (e.g., free-text responses), addressing the following topics:

- Demographic information: Age, sex, country of employment, level of education, primary area of expertise, publication record, and years of experience as a peer reviewer.
- Experience with AICs: Participants’ familiarity with AICs, prior use of AICs in their professional or academic work, and likelihood of permitted AIC use in the peer review process in the future.
- Role of AICs in peer review: Perceptions regarding the potential roles of AICs in peer review, such as aiding in identifying methodological flaws, detecting plagiarism, verifying references, translating research materials, or assessing the quality of writing.
- Perceived benefits of AICs in peer review: Participants’ views on potential benefits, such as reducing workload, improving efficiency, standardizing review quality, addressing biases, and increasing consistency in decision-making.
- Perceived challenges of AICs in peer review: Concerns about the reliability and accuracy of AICs, issues with authorship of peer review reports, risk of amplifying biases, lack of transparency, and potential over-reliance on AI.
- Ethical considerations: Participants’ perceptions of the ethical implications of integrating AICs in peer review, including concerns about accountability, confidentiality, data privacy, intellectual property rights, and their potential impact on the integrity of the peer review process.
- Open-ended questions: Participants had the opportunity to provide additional comments and feedback on the use of AICs in peer review, share their opinions on future integration, and suggest potential areas for improvement or guidelines for ethical use.

### Data Management and Analysis

All responses were collected via SurveyMonkey, and the data was exported and analyzed using Microsoft Excel 2007. Descriptive statistics, such as frequencies and percentages, were calculated to summarize the survey response data. Demographic information, such as participant role and years of experience, was analyzed for trends or patterns and described. Qualitative data collected through open-ended questions underwent inductive coding and thematic analysis by two authors (DB and ND) [29, 30]. To ensure consistency in coding, each author independently coded the responses of the first three survey responses for question 16 and then collaborated to develop a shared codebook based on their results. After developing a consensus on the code(s) and a unified coding framework, all remaining open-ended responses were coded, and the codebook was iteratively updated. Following this, individual codes were grouped into the themes by the two authors independently and finalized through consensus. Any conflicts were resolved through discussion between the two authors and if needed, by a third author (JYN). The two authors (DB and ND) then developed a clear definition for each theme based on codes incorporated. All data was then reviewed by JYN, DB, and ND. The final manuscript was written in accordance with the Strengthening the Reporting of Observational Studies in Epidemiology (STROBE) and the Checklist for Reporting Results of Internet E-Surveys (CHERRIES) guidelines [31,32].

## Results

Our collected raw survey data from SurveyMonkey can be found in **Appendix 3** (https://osf.io/varje/files/wq98x). Any personal identifiers provided by participants have been redacted. Additionally, crosstabs for key demographic variables (age, career stage, researcher vs. clinician, sex, and WHO regions) can be found in **Appendix 4** (https://osf.io/varje/files/c94uk).

### Respondent Demographics

Survey invitations were sent to a total of 72,851 email addresses, of which 1260 invitees provided responses. However, of the 72,851 email invites, there were 6,172 bounced emails (response rate: 1.9% or 1260/66,679) and an additional 32,839 emails that remained unopened by the invitee. Hence, only 33,840 email invites were received and opened by potential respondents (response rate: 3.7% or 1260/33,840). Of the respondents, 1157 met the eligibility criteria (91.8%), of whom 1077 completed the survey (93.1% completion rate). Among those that completed all survey questions (∼844/1260 or ∼67%), the average completion time was approximately 24 minutes. As not all respondents completed all survey questions, the total number of responses for each question varies and is provided in parentheses throughout the presentation of results that follow.

Demographic data is shown in **Table 1**. The majority of respondents identified as male (n=592 of 1077, 55.0%) and 19 respondents indicated ‘other’ or preferred not to disclose (1.8%). Respondents were employed across the six World Health Organization World Regions, with the greatest representation from the Americas (n=416/1077, 38.6%), followed by Europe (n=377, 35.0%) and South-East Asia (n=111, 10.3%). Approximately half of the respondents stated that English was not their primary language (n=581/1077, 54.0%).

**Table 1:**
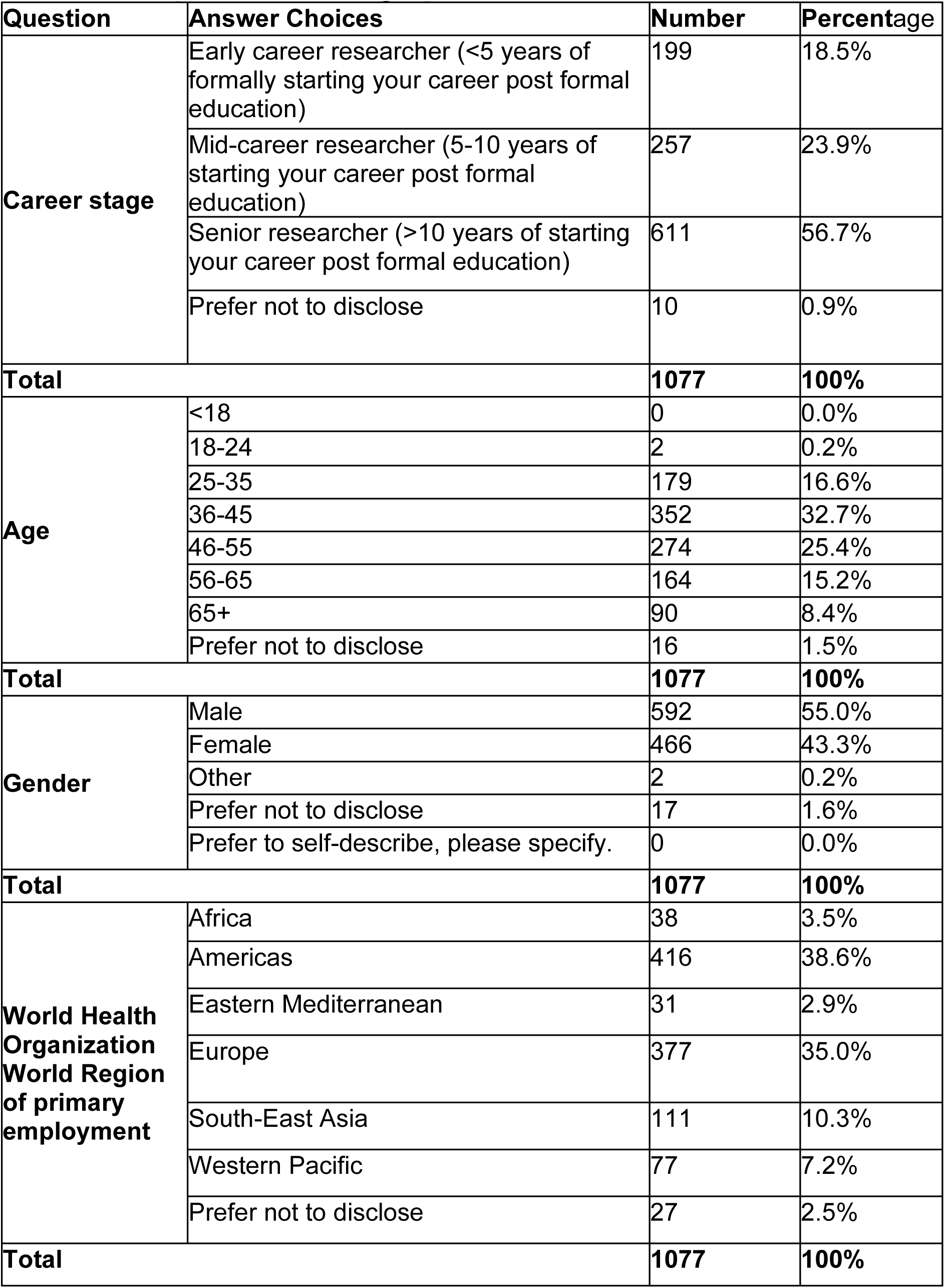
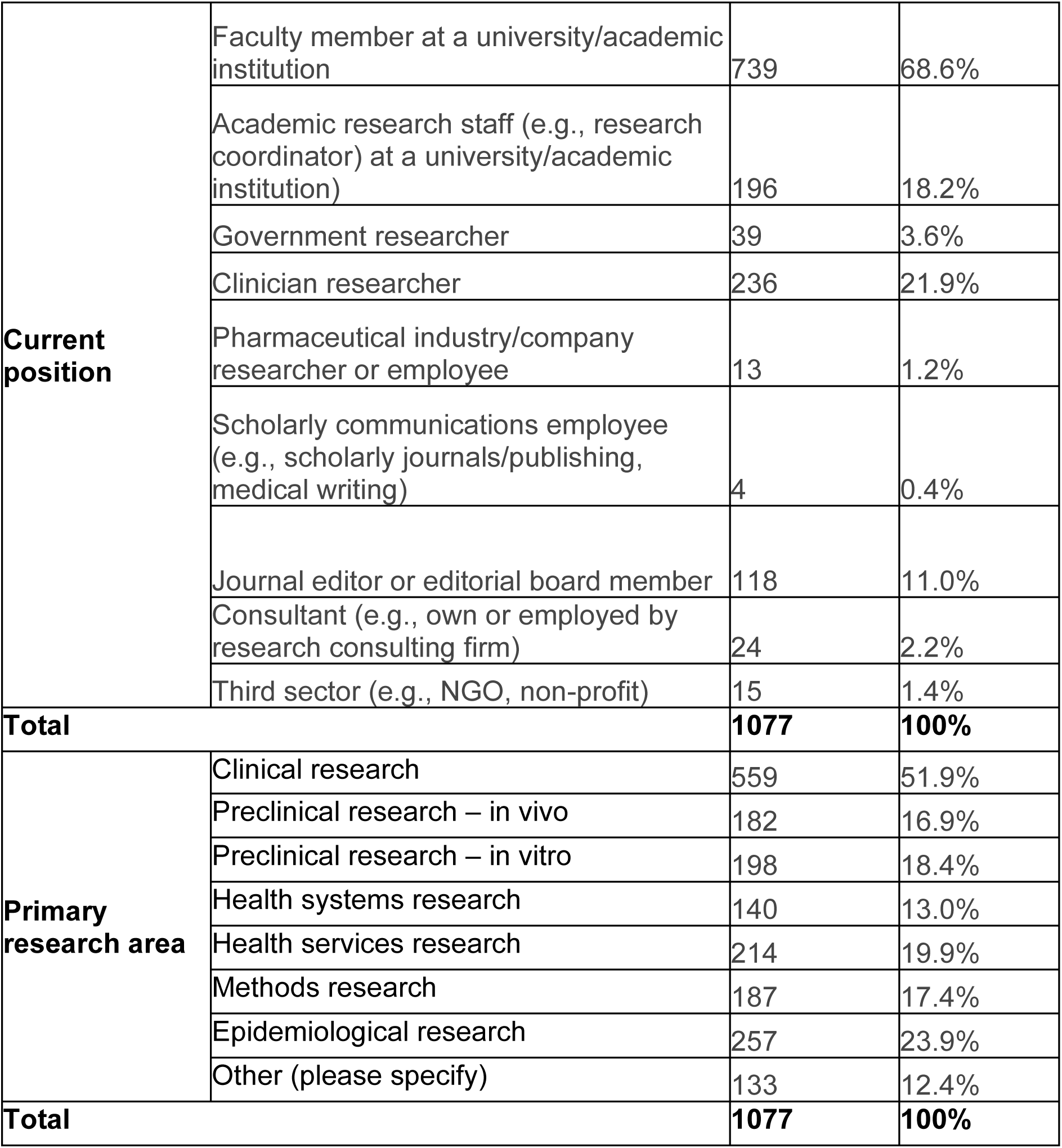
Respondent Demographic Data.

| Question | Answer Choices | Number | Percentage |
| --- | --- | --- | --- |
| **Career stage** | Early career researcher (<5 years of formally starting your career post formal education) | 199 | 18.5% |
|  | Mid-career researcher (5-10 years of starting your career post formal education) | 257 | 23.9% |
|  | Senior researcher (>10 years of starting your career post formal education) | 611 | 56.7% |
|  | Prefer not to disclose | 10 | 0.9% |
| **Total** |  | **1077** | **100%** |
| **Age** | <18 | 0 | 0.0% |
|  | 18-24 | 2 | 0.2% |
|  | 25-35 | 179 | 16.6% |
|  | 36-45 | 352 | 32.7% |
|  | 46-55 | 274 | 25.4% |
|  | 56-65 | 164 | 15.2% |
|  | 65+ | 90 | 8.4% |
|  | Prefer not to disclose | 16 | 1.5% |
| **Total** |  | **1077** | **100%** |
| **Gender** | Male | 592 | 55.0% |
|  | Female | 466 | 43.3% |
|  | Other | 2 | 0.2% |
|  | Prefer not to disclose | 17 | 1.6% |
|  | Prefer to self-describe, please specify. | 0 | 0.0% |
| **Total** |  | **1077** | **100%** |
| **World Health Organization World Region of primary employment** | Africa | 38 | 3.5% |
|  | Americas | 416 | 38.6% |
|  | Eastern Mediterranean | 31 | 2.9% |
|  | Europe | 377 | 35.0% |
|  | South-East Asia | 111 | 10.3% |
|  | Western Pacific | 77 | 7.2% |
|  | Prefer not to disclose | 27 | 2.5% |
| **Total** |  | **1077** | **100%** |
| **Current position** | Faculty member at a university/academic institution | 739 | 68.6% |
|  | Academic research staff (e.g., research coordinator) at a university/academic institution) | 196 | 18.2% |
|  | Government researcher | 39 | 3.6% |
|  | Clinician researcher | 236 | 21.9% |
|  | Pharmaceutical industry/company researcher or employee | 13 | 1.2% |
|  | Scholarly communications employee (e.g., scholarly journals/publishing, medical writing) | 4 | 0.4% |
|  | Journal editor or editorial board member | 118 | 11.0% |
|  | Consultant (e.g., own or employed by research consulting firm) | 24 | 2.2% |
|  | Third sector (e.g., NGO, non-profit) | 15 | 1.4% |
| **Total** |  | **1077** | **100%** |
| **Primary research area** | Clinical research | 559 | 51.9% |
|  | Preclinical research – in vivo | 182 | 16.9% |
|  | Preclinical research – in vitro | 198 | 18.4% |
|  | Health systems research | 140 | 13.0% |
|  | Health services research | 214 | 19.9% |
|  | Methods research | 187 | 17.4% |
|  | Epidemiological research | 257 | 23.9% |
|  | Other (please specify) | 133 | 12.4% |
| **Total** |  | **1077** | **100%** |

Many respondents were senior researchers (n=611/1077, 56.7%), faculty members at a university/academic institution (n=739/1077, 68.6%), primarily conducted clinical research (n=559/1077, 51.9%), published more than 50 peer-reviewed research articles (587/1077, 54.5%) and peer-reviewed more than 50 research articles (560/1077, 52.0%). Nearly 1 in 5 (n=199/1077, 18.5%) were also early-career researchers with less than 5 years of formal career experience in scholarly publishing.

### Experience with AICs

**Table 2** provides the full details on respondent experiences with AICs and their perspectives on incorporating AICs into the peer review process. Most respondents (n=917/1064, 86.2%) were familiar with the concept of AICs, and 25.6% (n=272/1064) stated they are very familiar with the concept. Only 2.8% (n=30/1064) were very unfamiliar with the concept. The majority had used ChatGPT (n=934/1065, 87.7%) for general purposes. When asked about which AICs they had used for purposes related to peer review, the majority of respondents indicated that they had never used an AIC for this purpose (n=744/1058, 70.3%). Of those who had used them for this purpose, 277 responses (26.2%) indicated ChatGPT. Respondents were either likely (n=278/1066, 26.1%) or very unlikely (n=282/1066, 26.5%) to use AICs for peer review purposes in the future.

**Table 2:**
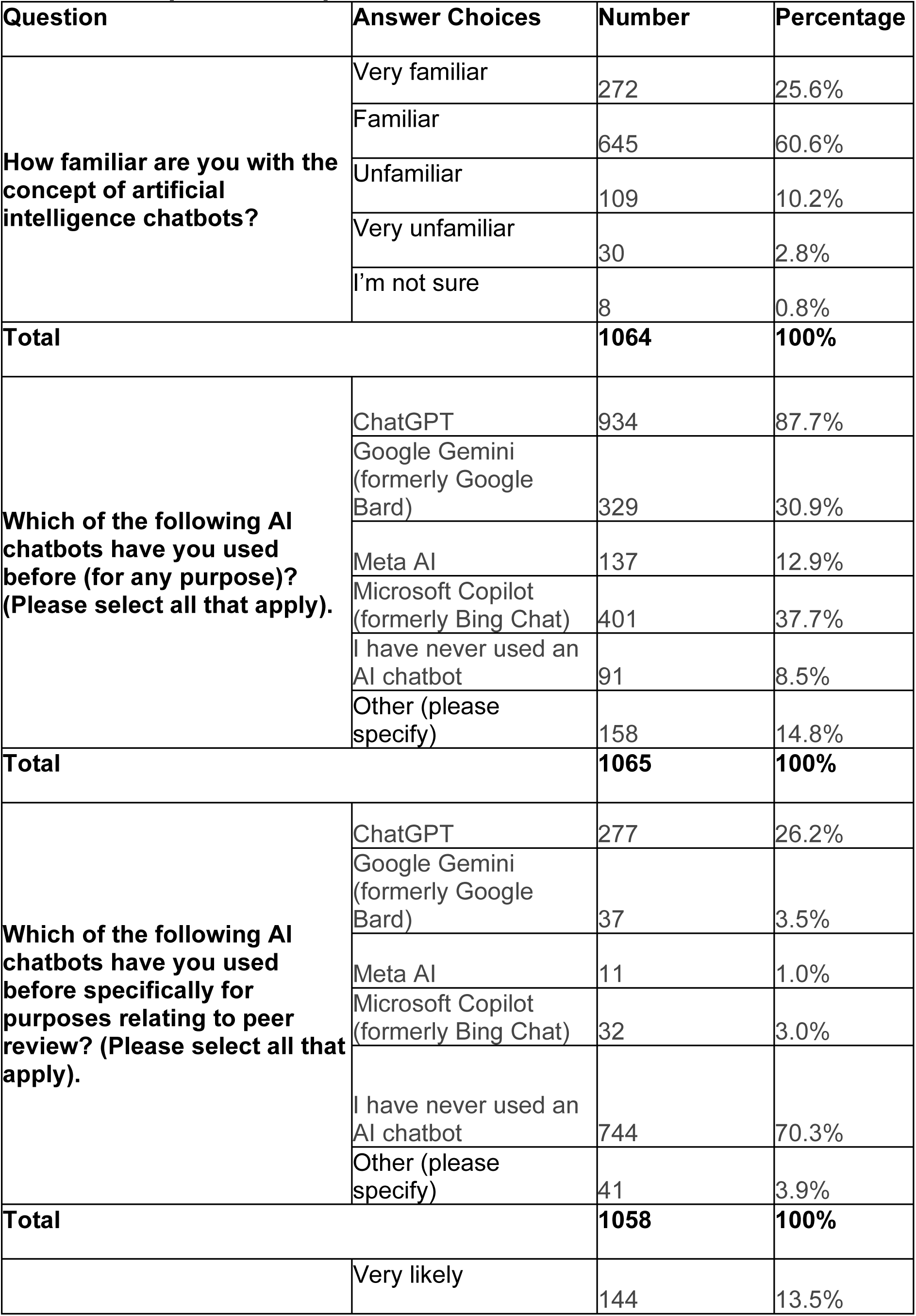
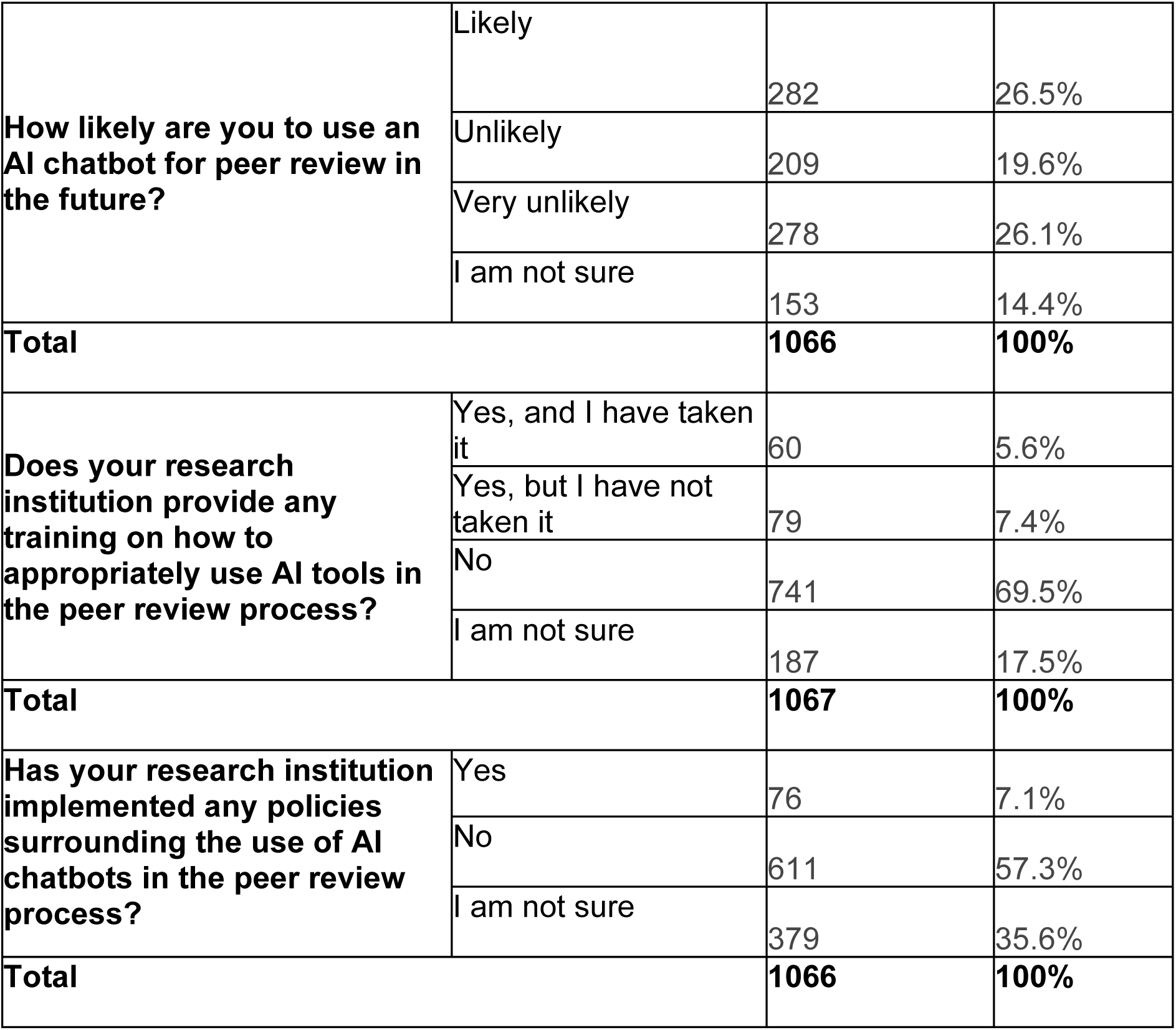
Respondent Experience with AI Chatbots.

Most respondents reported that their research institution does not provide any training on using AICs appropriately in the peer review process (n=741/1067, 69.5%). Some (139/1067,13.0%) reported that their institution did provide this training, however, only 60/139 (43.2%) had taken this training. Over half of the respondents indicated their research institution had not implemented any policies surrounding the use of AICs in the peer review process (n=611/1036, 57.3%), while only 76/1066 (7.1%) said that their institution did implement these types of policies.

### Role of AICs in Different Steps of the Peer Review Process

All responses regarding how they perceive the role of AICs in the peer review process are shown in **Table 3**. The respondents were shown a list of 12 different steps of the peer review process and asked to indicate how often they have used AICs to assist them with each step, on a 5-point scale ranging from “Always” to “Never” (**Figure 1)**. Across all steps, the majority of the respondents stated that they had never used AICs to assist them in peer review. Notable findings include that 90.3% (n=933/1033) stated that they had never used AICs to assist them with identifying potential conflicts of interest, while 1.0% (n=10/1033) indicated that they always used AICs to support them for this purpose. Of the respondents, 87/1039 (8.4%) had always or often used AICs to summarize the main research question and findings of a manuscript, 158 (15.2%) had sometimes or rarely used AICs for this purpose, and 794 (76.4%) had never used them for this purpose. Regarding providing feedback to the authors, 755/1036 (72.9%) indicated that they had never used AICs for this purpose, 165 (15.9%) had sometimes or rarely used them, and 116 (11.2%) had often or always used them.

**Figure 1:**
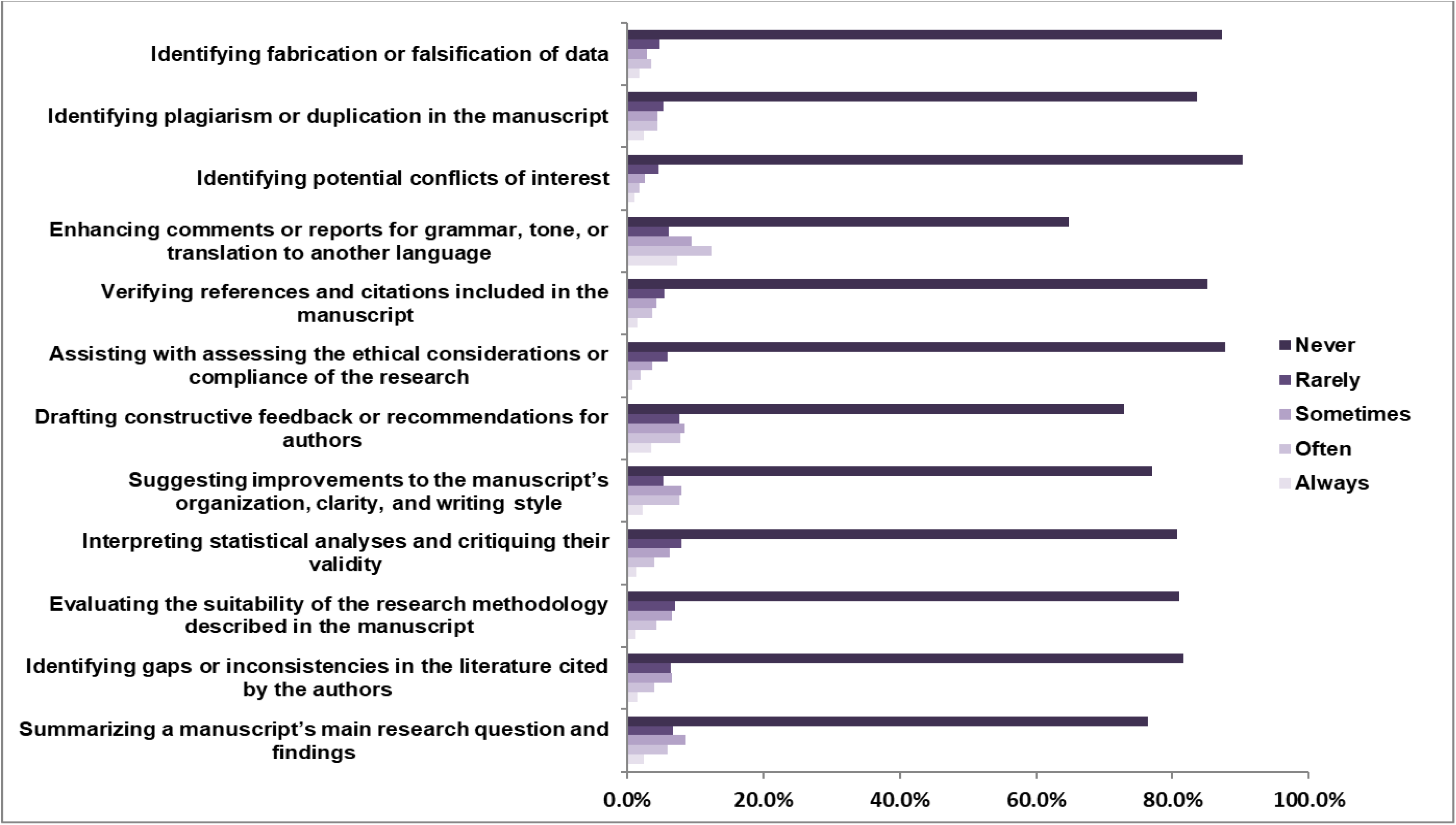
Respondent Use of AI Chatbots in Different Steps of the Peer Review Process

**Table 3:** Role of AI Chatbots in the Peer Review Process.

| Question | Answer Choices | Number | Percentage |
| --- | --- | --- | --- |
| **How much training and education do you think researchers need in order to effectively use AI chatbots in the peer review process?** | A lot | 396 | 38.3% |
|  | Some | 492 | 47.5% |
|  | Very little | 63 | 6.1% |
|  | None | 23 | 2.2% |
|  | I am not sure | 61 | 5.9% |
|  | **Total** | **1035** | **100%** |
| **Would you be interested in learning more/receiving training about how to use AI chatbots in the peer review process?** | Yes | 629 | 60.7% |
|  | No | 166 | 16.0% |
|  | Maybe | 241 | 23.3% |
|  | **Total** | **1036** | **100%** |
| **How important do you think AI chatbots will be in the future of peer review?** | Very important | 311 | 30.1% |
|  | Important | 441 | 42.6% |
|  | Unimportant | 72 | 7.0% |
|  | Very unimportant | 43 | 4.2% |
|  | I'm not sure | 168 | 16.2% |
|  | **Total** | **1035** | **100%** |
| **In general, how do you perceive the potential impact of AI chatbots on the future of peer review?** | Very positively | 139 | 13.4% |
|  | Positively | 419 | 40.4% |
|  | Negatively | 185 | 17.9% |
|  | Very negatively | 100 | 9.7% |
|  | I am not sure | 193 | 18.6% |
|  | **Total** | **1036** | **100%** |

When asked about how much training and education a researcher would need to effectively use AICs for peer review, 492/1035 respondents (47.5%) answered that ‘some’ training is necessary, while 396 (38.3%) said that ‘a lot’ of training was necessary. Most respondents expressed that they would be interested in learning more/receiving training on using AICs in the peer review process (n=629/1036, 60.7%). Most respondents felt that AICs would be ‘very important’ (n=311/1035, 30.1%) or ‘important’ (n=441, 42.6%) in the future of peer review.

Many respondents reported that AICs would have a ‘very positive’ or ‘positive’ outcome (n=558/1036, 53.9%) on the future of peer review, while 285 (27.5%) indicated AICs would have a ‘negative’ or ‘very negative’ impact, and 193 (18.6%) were ‘not sure’ how AICs would impact future peer review.

### Perceived Benefits and Challenges of AI Chatbots in the Peer Review Process

Respondents rated their agreement with statements relating to how beneficial AICs could be in different steps of the peer review, on a 5-point scale ranging from “Strongly disagree” to “Strongly agree” (**Figure 2)**. The most common benefits of AICs included enhanced quality, clarity and readability of peer review reports by suggesting edits to grammar, language, tone, or structure (n=652/1013, 64.4%); reduced workload for peer reviewers (n=634/1014, 62.5%); increased speed and efficiency of conducting reviews (n=586/1014, 57.8%); enabling reviewers to detect potential instances of plagiarism or duplication (n=583/1010, 57.7%); and generating constructive feedback, comments, or recommendations for authors (n=518/1011, 51.2%).

**Figure 2:**
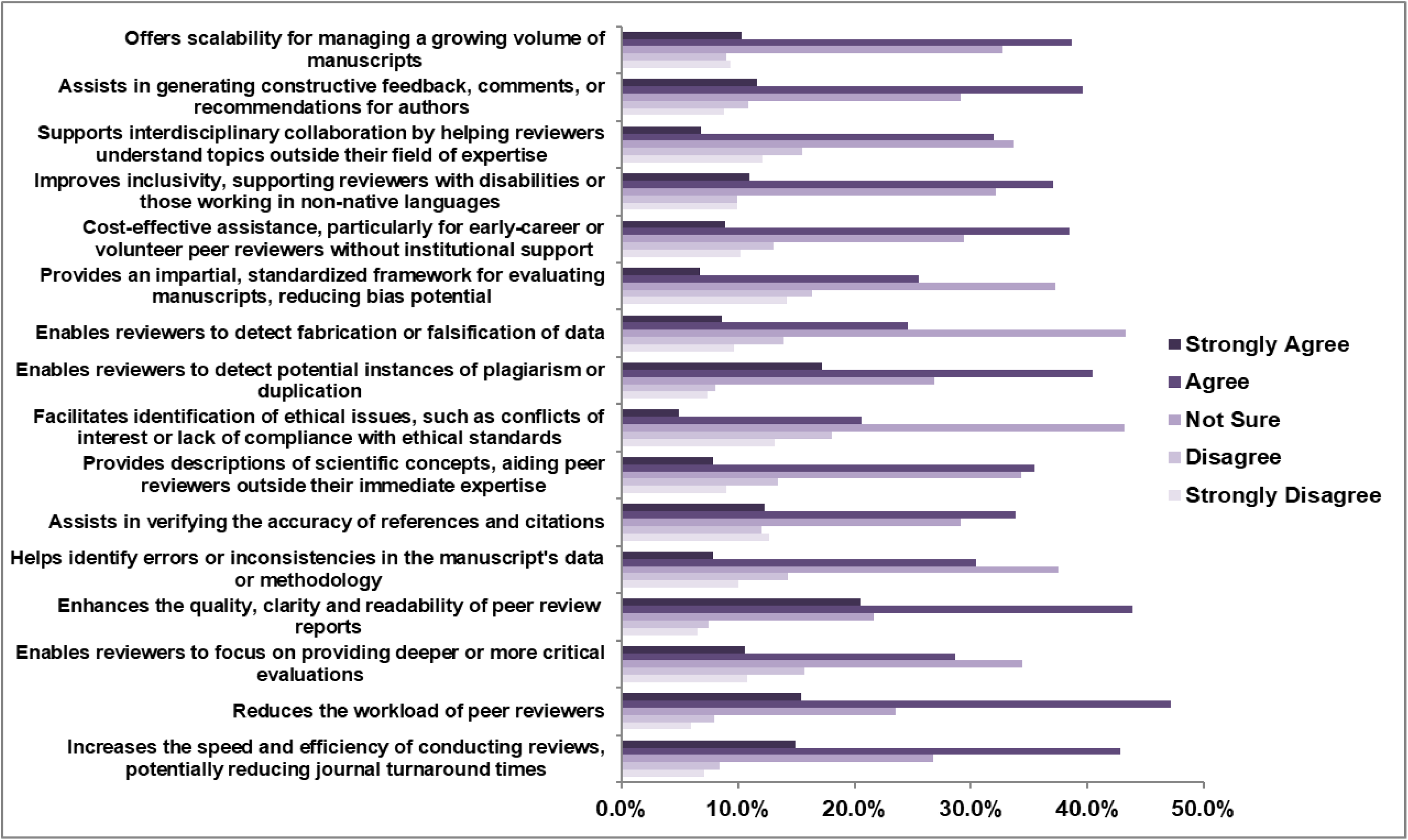
Respondent Agreement with Potential Benefits of Using AI Chatbots in the Peer Review Process

There was a mixed response about whether AICs could allow reviewers to focus on providing deeper or more critical evaluations. 268/1014 (26.4%) respondents strongly disagreed or disagreed with the statement, 397 (39.2%) responses strongly agreed or agreed, and 349 (34.4%) were not sure. Also, 379/1011 (37.5%) were unsure if AICs could help identify errors or inconsistencies in the data or methodology of a manuscript, while 387 (38.3%) strongly agreed or agreed with the statement, and 245 (24.2%) strongly disagreed or disagreed with the statement.

Respondent views were mixed on whether AICs could assist with the detection of fabrication or falsification of data. Respondent views were also mixed on whether AICs could provide an impartial and standardized framework for evaluating manuscripts, reducing the potential for reviewer bias as 143/1010 (14.2%) strongly disagreed, 165 (16.3%) disagreed, 376 (37.2%) were unsure, 258 (25.5%) agreed and 68 (6.7%) strongly agreed.

The respondents rated their agreement with statements relating to potential challenges of using AICs in the peer review process on a 5-point scale ranging from “strongly disagree” to “strongly agree” (**Figure 3**). The most agreed upon challenges of AICs included biased or skewed evaluations due to AIC algorithm limitations (n=794/989, 80.3%), risks of producing errors or inaccuracies due to limited comprehension of jargon and limitations inherent to the AICs (n=784/989, 79.3%), requiring continuous updates and training to remain effective with advancements in scientific methods and emerging topics (n=777/986, 78.8%), struggles with the complexity and nuance of interpreting scientific arguments and methodologies in manuscripts (n=756/989, 76.4%), and a risk in oversimplifying or overlooking subtleties in arguments or findings (n=754/985, 76.5%).

**Figure 3:**
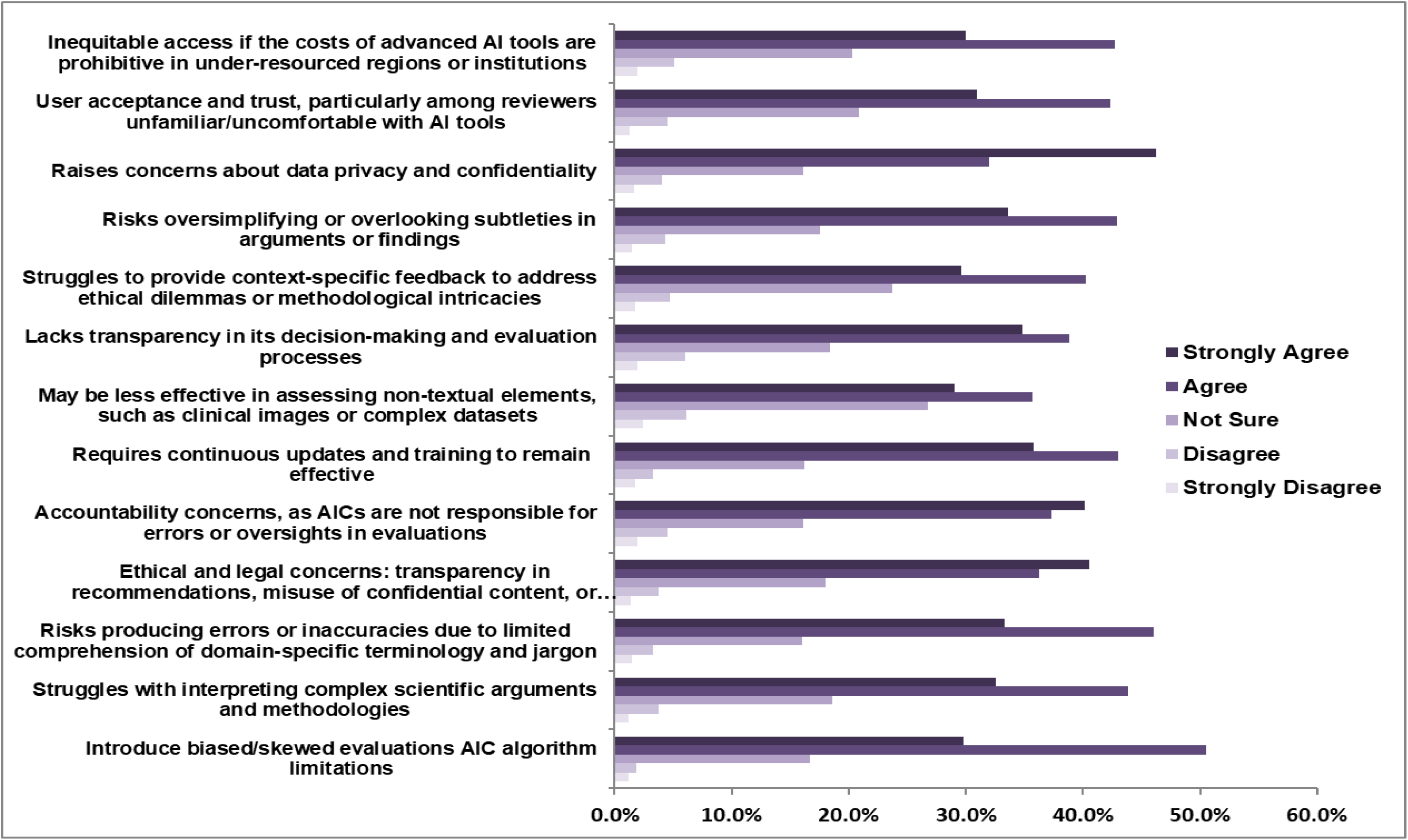
Respondent Agreement with Potential Challenges of Using AI Chatbots in the Peer Review Process

### Open-ended Questions

The six open-ended questions related to training, policies, use, benefits, and challenges, allowed participants to provide additional feedback and comments on the use of AICs in peer review. Across all questions and responses, 25 themes and 83 codes were identified. Each response was annotated with only one code; each code was grouped under only one designated theme. Recurring themes included appreciation of AIC roles in peer review processes for ‘proofreading’, ‘learning [manuscript] topics’, ‘content verification’, and publisher tools such as ‘translation’ and ‘statistical analysis’ as many believe AICs are “increasing” opportunities to publish and review diverse research”. However, peer reviewers also endorsed common concerns: “issues with hallucinations”, ‘overautomation of peer review processes’ and ‘compromised research quality assurance’. AICs may also lack the ability to provide context-specific feedback of novel, complex manuscript content as respondents state that “feedback from chatbots can be too general” and “present harmful critique of transformative new ideas”. Altogether, responses were about evenly-divided in positive and negative sentiments of AIC use in peer review processes. Broad perspectives included either a ‘favourable view of AI adoption’ (e.g., “helps a lot getting an overview of the work”, “a very good first approach”) and ‘acceptance with caution’, or firm opposition with many echoing that “using [AICs] for peer review is not an appropriate shortcut”, emphasizing the above-mentioned concerns and others such as confidentiality concerns (e.g., “tantamount to theft…copyrights may not be protected”)” and “lack of training [being] a big hurdle” that threaten the integrity of scholarly publishing in medical journals. The themes and code frequencies can be found in **Appendix 5** (https://osf.io/varje/files/cehpj). Full results to the thematic analysis tallies can be found in **Appendix 6** (https://osf.io/varje/files/u3ypj). A supplemental quantitative analysis of assigned codes for each question can be found in **Appendix 7** (https://osf.io/varje/files/23h4y).

## Discussion

### Significance of Findings

This survey is one of the first of its size and scope to offer a global perspective on the attitudes and perceptions of medical journal peer reviewers regarding the use of AICs in the peer review process, and their potential impact on how peer review is conducted at a time when these AICs are being increasingly applied to scholarly publishing.

Our survey found that the majority of respondents were already familiar with the concept of an AIC and had utilized AICs before for general purposes, with ChatGPT being the most commonly used AIC. However, most had indicated that they had not used AICs for any purpose related to peer review, specifically. They expressed diverging views regarding their future use of AICs in peer review, with most indicating they were either likely or highly unlikely to use AICs for this purpose.

Respondents also held mixed views on the potential benefits of AICs for peer review, though there was more consensus regarding the limitations and disadvantages associated with their use. Respondents expressed that while AICs have potential to support peer review and the various steps within the process, they present with many challenges and errors pertaining to their use that need to be considered.

Most respondents reported that their research institution did not provide training on the appropriate use of AICs in the peer review process. Only 139 respondents reported that their institution did provide this training; however, only about half of these respondents had participated in the training. Additionally, very few had reported that their institutions implemented policies surrounding AIC use in peer review. This lack of training and policies surrounding AIC use in peer review, and scholarly publishing in general, could be due to limited awareness or adoption of these tools, especially as many publishing organizations and journals have stated that AI tools cannot be used for research publications [33]. Given that most peer reviewers in our survey expressed that they do not use AICs for peer review, it could be that there was a lack of perceived need for these types of training and policies.

Similarly, these resources may be lacking as they may not be considered core academic responsibilities of research institutions and academic publishers, but rather voluntary tasks that must be undertaken by scholars. However, a small number of participants did indicate that they used AICs for peer review, and most participants stated that reviewers did need training and education to effectively use AICs for peer review with most indicating interest in receiving this training. Additionally, the discussion surrounding the use of AICs in scholarly publishing has raised several concerns, including ethical issues, risks of bias, and challenges related to the accuracy and transparency of automated responses, leading many to call for transparency and education regarding their “responsible” use throughout the publishing processes [34].

Recent studies have raised well-supported concerns regarding the integrity of research involving AIC use due to the lack of action by many academic journals in implementing robust guidelines and controls for AI-generated content [35]. In addition, unreported use and overreliance of AICs has the potential to decrease the quality of peer review, which in turn impacts the credibility of research being published [36]. AICs may lack the contextual understanding and nuance needed to effectively assess research methodology and framing. The consequences of this deficit may include missed errors or inadequate assessment of key aspects of a manuscript, especially for novel research that requires in-depth, context-specific expertise to provide a thorough evaluation [36]. Therefore, without robust quality control measures implemented for AIC use in peer review and publishing, there is significant cause for concern regarding the credibility and integrity of research.

However, this limitation may be addressed by assigning responsibility for quality control to the editorial office or academic publisher rather than individual peer reviewers.

### Comparative Literature

Although the potential role of AIC use in the peer review process is a widely discussed topic in the medical scholarly publishing community, there are very few similar studies that have directly explored the attitudes and perceptions of editors and peer reviewers [25, 37–40]. To consider the incorporation of AICs into peer review, it was important to gauge the standpoints of these potential users. Hence, studies incorporating questionnaires, interviews, or surveys were considered comparative literature.

For example, a related study involving a similar international cross-sectional survey of 2165 corresponding authors was conducted in 2023 to understand the attitudes and perceptions of medical researchers toward AIC use in the scholarly publishing process, in general [37]. Most authors perceived a positive potential impact of AICs on the future of scientific research (1236/2048 or 60.4%) and stated that they were likely to use them for upcoming research (1241/2136 or 58.1%). The majority emphasized that training and education would be needed for the effective use of AICs (1713/2049 or 83.7%) but reported that training was not offered by their research institution (1487/2137 or 69.6%) [37]. The study concluded from the survey findings that supportive educational initiatives are warranted for the increasing body of researchers familiar and experienced with AIC use in scientific research.

Similarly, an international survey conducted by the academic publisher *Springer Nature* in March 2025 explored the viewpoints of 5229 researchers on the role of AI tools in the peer review processes of scholarly publishing, non-specific to any discipline [25]. Although approximately 90% of the respondents reported appropriateness of AI use for purposes such as proofreading and translation, with or without disclosure, only 34% of the respondents agreed that it was appropriate to use an AI tool to create an initial peer review report. The majority (52%) stated that using AI to create a peer review report was inappropriate “under any circumstances”, whereas 14% cited inappropriateness “because of privacy concerns” [25]. The majority (57%) also reported that the use of AI tools by peer reviewers to fact-check or clarify questions regarding a specific manuscript (e.g., correct use of a statistical technique) was appropriate. Yet, most respondents claimed that they had neither used AI tools, or would even consider their use, in peer review for initial peer review (78%) or clarifying questions (56%) [25].

Another related international cross-sectional survey, conducted in 2024, exclusively explored the perspectives of 510 Editors-in-Chiefs (EiCs) of biomedical journals regarding the use of AICs in scholarly publishing [38]. Similar to the findings of this survey and other studies [25, 37], the majority of EiCs had not used AICs for editorial purposes (401/479 or 83.7%) though expressed interest in receiving training (302/469 or 64.4%). Overall, the study reported mixed attitudes as many EiCs would acknowledge benefits of AICs, such as enhancing proofreading abilities, whilst raising concerns about initial resources required for training and education and ethical implications of data privacy [38]. Hence, the need for clear guidelines regarding responsible AIC use is again emphasized, in addition to longitudinal assessments of the impact of AIC adoption on the quality and integrity of the editorial or peer review process.

Two additional studies of similar design and purpose surveyed 17 peer reviewers and editors, respectively, of unspecified fields and disciplines, to gauge their general attitudes toward the role of AICs in editing and peer review [39, 40]. Their results were similar as most participants expressed mixed opinions: AICs can enhance the readability and efficiency of peer review reports, but lack context-based feedback for authors. Both studies also emphasize that AICs in peer review threaten the essential “human touch”, and call for guidelines that ensure responsible use with continued human involvement [39, 40]. Descriptive statistics were not reported.

Finally, the two most recent studies following the administration of our survey were conducted by academic publishers *Frontiers* and *IOP Publishing* [41, 42]. A global reviewer survey administered by *IOP Publishing* in August 2025 gauged the viewpoints of 348 physical science researchers on the uses of generative AI in peer review processes [41]. Although 32% of respondents admitted to using AI tools for support with peer review tasks, and 40% of respondents perceived a positive impact of AI for peer review, many expressed dissatisfaction regarding the use of AI tools to write (∼57%) or augment (∼42%) a peer review report for their submitted manuscript [41]. Similarly, a global survey of 1645 active researchers in June 2025 *Frontiers* found that approximately 53% of reviewers use AI tools in peer review processes, largely by early career researchers (∼87%) for synthesizing peer review reports [42]. However, only 21% of respondents agreed that AI use “increases their trust in publishers” with 45% having concerns of misuse [42]. Therefore, both publishers have concluded from their respective studies that although there is an accelerated adoption of AI use by peer reviewers, there remains an urgent need for policy development that can responsibly nurture the potential of AI use for peer review without compromising scientific rigor or research integrity [41, 42].

### Future Directions

There are concerns about the use of AICs undermining the need for rigorous, unbiased, and transparent review practices. To guide the responsible integration of AICs in peer review, a clear ethical framework is necessary. Future research should focus on developing these frameworks to allow peer reviewers to better understand these tools and how they can be used to support peer review while mitigating harmful consequences of their use. This could inform the development of academic journal or publisher guidelines for the responsible use of AICs in the peer review process, balancing their potential benefits with the need for rigorous, unbiased, and transparent review practices. Additionally, developers would need to address the limitations of AICs and ensure their continuous improvement to minimize errors, such as hallucinations, and substandard, generic output. Once these pertinent limitations have been addressed, a cross-sectional survey of a similar design may be repeated to help gauge possible changes in peer reviewer perspectives on the role of AICs, if any, in the peer review process.

### Strengths and Limitations

This study used a cross-sectional survey design, which offers several strengths and limitations. One of the key strengths is that this survey approach is cost-effective. As the study only observes a specific population at a single point in time, it was quick to administer and gather data from a large sample of medical journal peer reviewers, allowing for the results to be generalizable to the broader medical research community. As the researchers in our sample are affiliated with diverse medical disciplines and WHO world regions, we received a broad range of opinions regarding the use of AICs in the peer review process, providing valuable insights. Additionally, by collecting names and email addresses of corresponding authors only from the previous two months, we minimized the likelihood of encountering inactive or bounced emails, which helped ensure the accuracy of our contact list.

There are also limitations to the design of this study. As with all cross-sectional surveys, there is the potential for recall bias, and selective non-response bias, which may impact the validity of our findings. Furthermore, self-reported attitudes and perceptions may be weak predictors of implicit biases and actual behaviour. For instance, there may be social desirability bias as peer reviewers may not accurately disclose AIC use for their tasks pertaining to peer review due to their concerns regarding professional and legal consequences, despite the anonymous nature of the survey. Another major limitation is that non-English speaking researchers are largely excluded from our sample due to language constraints, which could affect the applicability of our findings to those who primarily publish in languages other than English. Similarly, researchers with limited English proficiency may have experienced difficulties in completing the survey, further narrowing the scope of our data. Moreover, while we made considerable efforts to encourage participation from all invitees, regardless of experience with AICs, the generalizability of our findings will likely be affected by response bias. Invitees with strong opinions for or against the use of AICs may have been more inclined to respond to our survey, and as such, those with limited experience or familiarity with AICs may be underrepresented in our sample. The overall response rate was relatively low, which further impacts generalizability. However, the final total number of participants nevertheless represent a relatively large sample size due to the scale of our sampling. Finally, the results from our study only provide a cross-sectional analysis of peer-reviewer perspectives. However, perspectives surrounding AIC use, and AIC uses themselves, are continuously evolving.

## Conclusion

This survey provides insight into peer reviewers’ attitudes and perceptions about AICs for peer review processes in medical journals. Many peer reviewers indicated in this survey that although they had not used AICs for peer review purposes, they were interested in learning more and receiving training on how to use them responsibly to reap the potential benefits in peer review processes. The majority also responded that their research institutes or affiliated journals did not have any training or policies in place regarding the use of AICs for peer review with many expressing concerns surrounding the limitations and ethical downfalls of AIC use medical journal peer review. Hence, given the growing interest, academic journals and publishers may look into developing available training opportunities and policies to help their peer reviewers understand the possible impacts of using AICs in their work of peer review. These steps would be crucial in helping journals responsibly incorporate AICs by preserving the rigour and integrity of peer review processes in medical scholarly publishing.

## Data Availability

All relevant study materials and data associated with this study have been made publicly available on the Open Science Framework.

https://doi.org/10.17605/OSF.IO/VARJE

## List of Abbreviations

AI: artificial intelligence
AIC: artificial intelligence chatbot
ChatGPT: Chat Generative Pre-Trained Transformer
COPE: Committee on Publication Ethics
LLM: large language model
PMID: PubMed identifier
OSF: Open Science Framework
WAME: World Association of Medical Editors

## Declarations

### Ethics Approval and Consent to Participate

Ethics approval was sought and obtained from the University Hospital Tübingen Research Ethics Board (REB Number: 080/2025BO2) to conduct this study.

### Consent for Publication

All authors consent to this manuscript’s publication.

### Availability of Study Materials and Data

All relevant study materials and data associated with this study have been made publicly available on the Open Science Framework, and can be found at https://doi.org/10.17605/OSF.IO/VARJE.

### Authors’ Contributions

JYN: designed and conceptualized the study, drafted the manuscript, and gave final approval of the version to be published.

DB: drafted and edited the manuscript and protocol, contributed to the study design, and gave final approval of the version to be published.

ND: drafted and edited the manuscript and protocol, contributed to the study design, and gave final approval of the version to be published.

LB: made critical revisions to the manuscript, contributed to the study design, commented on the study protocol, and gave final approval of the version to be published.

TC: made critical revisions to the manuscript, contributed to the study design, commented on the study protocol, and gave final approval of the version to be published.

HC: made critical revisions to the manuscript, contributed to the study design, commented on the study protocol, and gave final approval of the version to be published.

AF: made critical revisions to the manuscript, contributed to the study design, commented on the study protocol, and gave final approval of the version to be published.

AI: made critical revisions to the manuscript, contributed to the study design, commented on the study protocol, and gave final approval of the version to be published.

CL: made critical revisions to the manuscript, contributed to the study design, commented on the study protocol, and gave final approval of the version to be published.

AM: made critical revisions to the manuscript, contributed to the study design, commented on the study protocol, and gave final approval of the version to be published.

DM: made critical revisions to the manuscript, contributed to the study design, commented on the study protocol, and gave final approval of the version to be published.

## Acknowledgements

We gratefully acknowledge our survey’s pilot testers. Those who consented to be acknowledged include Luka Ursić, Wael Abdelkader, Rita Jezrawi, and Marija Franka Zuljevic. We also gratefully acknowledge Daniel Fry and Wid Al-Zahraa Al-Khafaji for their efforts in retrieving and exporting the PMID numbers of yielded MEDLINE-indexed journal articles to extract the contact information of our survey invitees.

